# Connectome-based Brain Marker Moderates the Relationship between Childhood Adversity and Transdiagnostic Psychopathology during Early Adolescence

**DOI:** 10.1101/2024.06.13.24308906

**Authors:** Xiang Xiao, Christopher J. Hammond, Betty Jo Salmeron, Danni Wang, Hong Gu, Tianye Zhai, Laura Murray, Annika Quam, Justine Hill, Hieu Nguyen, Hanbing Lu, Elizabeth A. Hoffman, Amy C. Janes, Thomas J. Ross, Yihong Yang

## Abstract

**Importance:** Identifying brain-based markers of resiliency that reliably predict who is and is not at elevated risk for developing psychopathology among children who experience adverse childhood experiences (ACEs) is important for improving our mechanistic understanding of these etiological links between child adversity and psychopathology and guiding precision medicine and prevention efforts for reducing psychiatric impact of ACEs.

**Objective:** To examine associations between ACEs and transdiagnostic psychopathology during the transition from preadolescence to early adolescence and test whether these associations are moderated by a hypothesized resilience factor, a previously identified connectome variate (CV) that is associated with higher cognitive function and lower psychopathology.

**Design, Setting, and Participants:** This study was conducted in a longitudinal design based on multicenter data from a community cohort of U.S. youth aged of 9-11 at baseline, who participated in the Adolescent Brain Cognitive Development (ABCD) study (N=7,382 at baseline and 6,813 at 2-year follow-up). Linear regression models and moderation analyses were used to characterize concurrent and prospective associations between lifetime ACEs and number of *DSM-5* psychiatric disorders (indexing transdiagnostic psychopathology) and to determine if individual variations in these associations were moderated by the CV derived from resting-state fMRI at baseline.

**Main Outcomes and Measures:** Cumulative number of current DSM-5 psychiatric disorders assessed using the computerized self-admin version Kiddie Schedule for Affective Disorders and Schizophrenia (KSADS-5) and lifetime ACEs assessed from child and parent reports at baseline (9-10 years) and 2-year-follow-up (11-12 years).

**Results:** ACE total scores correlated positively with the cumulative number of current DSM-5 psychiatric disorders at both baseline (*r* =.258, *p* < .001) and 2-year follow-up (*r* =.257, *p* < .001). The baseline CV score moderated the ACE-disorder associations at baseline (B = -0.021, *p* < .001) and at 2-year follow-up (B = -0.018, *p* = .008), as well as the association between the changes in ACE and in the number of disorders from baseline to year 2 (B = -0.012, *p* = .045). Post-hoc analyses further showed that the moderation effect of CV on ACE-psychopathology associations was specific to the threat-related ACEs and to female youth.

**Conclusions and Relevance:** These findings provide preliminary evidence for a connectome-based resiliency marker and suggest that functional connectivity strength in a broad system including frontal-parietal cortices and subcortical nuclei relevant to cognitive control may protect preadolescents who have experienced lifetime ACEs--especially females and those experiencing threat-related ACEs--from developing transdiagnostic psychopathology.

## Introduction

Adverse experiences in childhood, such as parental mental illness or witnessing violence in the community, are common among children living in the United States (U.S.).^1^ More than half of U.S. adolescents and adults report experiencing at least one type of adverse childhood experience (ACE) before age 18 and nearly 1-in-6 report experiencing 4 or more types of ACEs.^2,3^ Childhood adversities are strongly and consistently associated with negative psychological health, physical health, academic/vocational achievement, and functional outcomes across the lifespan.^4–6^ Regarding psychological health, ACEs appear to increase the risk for developing psychopathology across a broad range of psychiatric diagnostic categories and hierarchical domains (e.g., internalizing problems, externalizing problems, and p-factor) pointing to potential transdiagnostic mechanisms.^2,7,8^

The neurobiological mechanisms through which childhood adversity contributes to risk for psychopathology (and conversely resiliency against this risk) are areas of intense research. Most of this work has been focused on risk mechanisms, with few studies conducted to date characterizing neurobiological mechanisms conferring resilience to ACEs. In this space, emerging evidence indicates that childhood adversity results in alterations in brain structure and function, hypothalamic-pituitary-adrenal gland (HPA) axis function and sympathetic/parasympathetic tone, inflammation, microbiome functioning, and markers of biological aging. ^6,9,10^ This has led researchers to hypothesize that childhood adversity becomes “biologically embedded” and, through these multi-systemic physiological changes, ACEs influence downstream health outcomes for exposed individuals.^9^ Recent studies indicate that some of the variance in outcomes following ACE exposure can be attributed to cumulative risk exposure based on the age-of-onset, number, and chronicity of adversity experienced across childhood.^11^ Sub-dimensions of ACEs may also play a role in heterogenous outcomes. For example, ACEs from threat versus deprivation-related experiences may lead to psychopathology through different intermediate pathways and mechanisms.^12^ Threat experiences, defined as exposures to experiences that involve harm or threat of harm to the child or a close other (e.g., physical abuse or domestic violence), are associated with atypical fear learning and emotion processing when controlling for deprivation experiences.^13^ In contrast, deprivation experiences, defined as the experience of reduced cognitive stimulation and social inputs in the environment (e.g., neglect), are associated with lower performance on cognitive tasks after controlling for threat exposure. Early evidence suggests that threat and deprivation experiences produce distinct patterns of dysregulation in brain function and structure.^14,15^ Through better understanding of how different types and cumulative risk exposure of ACEs result in differential risk for psychopathology, we can improve prevention strategies for these youth.

Despite ACEs being a strong predictor of psychopathology, the relationships between ACEs and psychopathology are not deterministic.^6^ Many youths who experience ACEs have healthy emotional adaptation to the stress produced by adversity, not developing psychiatric problems as a consequence. Certain intrinsic processes and environmental factors (termed “resiliency factors”) may buffer children from risk for negative outcomes following ACEs.^16–18^ The neurobiological mechanisms underlying resilience in the face of childhood adversity are poorly understood but hypothesized to involve circuits engaged during cognitive control of emotions.^18–21^ Identifying brain mechanisms that confer intrinsic resilience and buffer against the development of psychopathology in youth who experience ACEs is critical for improving our mechanistic understanding of the etiological links between ACEs and psychopathology and can be used to enhance early identification and prevention efforts for vulnerable youth.

Neuroimaging techniques such as resting-state functional magnetic resonance imaging (rsfMRI) and resting-state functional connectivity (rsFC) analysis, enable noninvasive investigation of the system-level organization of brain circuits via the temporal synchrony between brain regions.^22^ When applied to the study of resiliency following childhood adversity, these methods can lead to better mechanistic understanding about who is and is not at risk for psychopathology and what brain systems are involved. The functional connectome, a collective set of functional connectivity (FC) across the whole brain, can reliably discriminate one brain from another like a fingerprint.^23^ Individual differences in functional connectome may underlie individual differences in cognitive and emotional processes relevant to resilience^23–25^, including development of psychiatric disorders.^26,27^ Our group recently applied machine learning to whole-brain rsFC data obtained from a rsfMRI scan and identified a connectome-based brain marker, mainly loaded in the fronto-parietal cortices and the subcortical system, that was positively associated with cognitive task performance, while negatively associated with psychopathology across domains. In the previous work, we found that the identified connectome variate (CV) predicted cumulative number of psychiatric diagnoses concurrently and prospectively in a population-representative sample of U.S. preadolescents ^28^.

To gain additional insights into mechanisms of resiliency and vulnerability to childhood adversity, the current study used longitudinal data from the Adolescent Brain Cognitive Development^(SM)^ (ABCD) study sample to investigate the impact of variation in this functional connectome patterns on concurrent and prospective associations between ACEs and transdiagnostic psychopathology during early adolescence in a large sample of preadolescents who vary in their exposure to ACEs. The aims of our study were three-fold: to test (1) whether ACEs are associated with the transdiagnostic psychopathology as measured by cumulative number of psychiatric disorders, (2) if the relationships between ACEs and transdiagnostic psychopathology are moderated by the CV that we have previously derived from whole-brain FC data, and, if a CV-moderating effect is shown, (3) if any such CV-moderating effects vary as a function of subdimension of ACE experienced (threat vs. deprivation) and sex.

## Method

### Participants and Study Design

Neuroimaging data, questionaries related to ACEs, and assessments for psychiatric disorders of 11,875 children aged 9-to 12-years were obtained from the ABCD study® ^29^. This large and long-term ongoing project aims to characterize psychological and neurobiological development from pre-adolescence through young adulthood. Participants and their families were recruited through school-based sampling frames at 21 centers across the U.S., following locally and centrally approved Institutional Review Board procedures as detailed elsewhere ^30^.

7,382 (3,714 females, aged 9.95 ± 0.62 y/o at baseline) of 11,875 participants met imaging analysis inclusion criterion for, detailed in our previous work.^28^ The functional connectome variate (CV) for each of the 7,382 participants was computed from the baseline resting-state fMRI ^28^. Clinical diagnoses of DSM-5 disorders were assessed using the computerized-version of the Kiddie Schedule for Affective Disorders and Schizophrenia for DSM-5 (KSADS-5), at both baseline and year-2 follow-up visits. ACEs at baseline and year-2 were derived from questionnaires collected within this period, based on the coding scheme in Stinson et al ^31^. Figure 1 shows the timeline of the datasets included in the current study, and the relevant variables and their pair-wise correlations.

**Figure 1.**
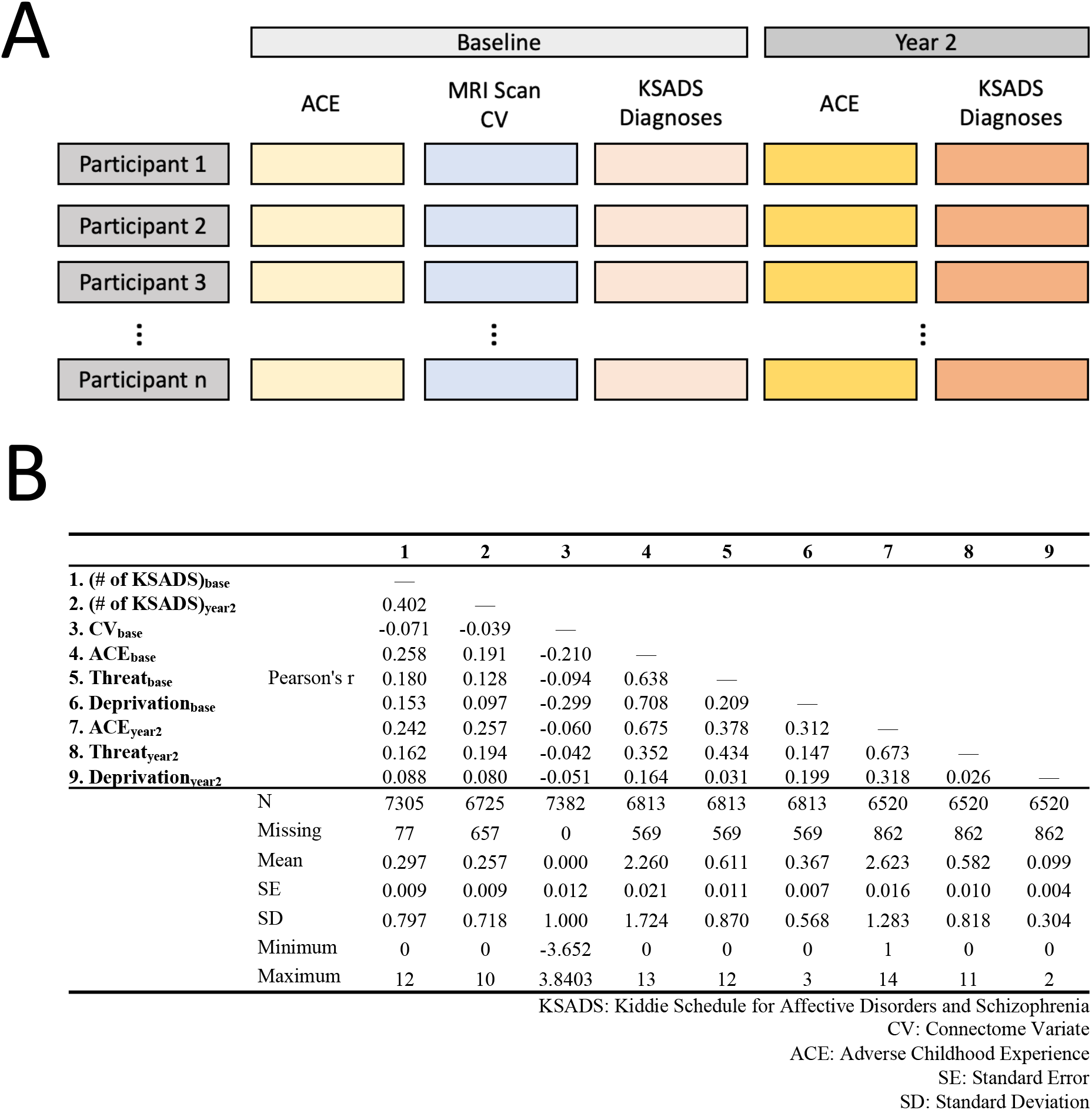
Study design and variables included in the analyses. A) Timeline of imaging and behavioral/clinical assessments. Connectome Variate (CV) was derived from the baseline functional MRI scan for each of individuals. Kiddie Schedule for Affective Disorders and Schizophrenia (KSADS) diagnoses, Adverse Childhood Experiences (ACEs) and its subdimensions, *Threat* and *Deprivation*, were assessed both at baseline and the 2-year follow-up. **B) Variables included in the analyses and their pair-wise correlations.** The upper part of the panel lists the pair-wise Pearson’s correlations among the 9 variables. The lower part of the panel lists the statistic summaries of each of the variables.

### Assessments of Adverse Childhood Experiences

ACEs are a set of potentially traumatic experiences occurring before adulthood, such as emotional and physical abuse, family dysfunction, parental neglect, and natural disasters, which can increase the risks of chronic physical and mental health conditions in adulthood. According to a previous study, ACEs in the ABCD study were scored by summing across children’s self-report and parent’s report of potentially traumatic events falling in ACE categories that youths have experienced in their lives ^31^. The ACE score can range between 0-21, with higher indicating greater severity. According to dimension model of ACE ^13^, two subdimensions, *Threat* and *Deprivation*, were derived from the ACE events. *Threat* includes 13 traumatic events reported in the PTSD survey and can range between 0-13. *Deprivation* includes 5 events, e.g., neglect and parental separation, and can range between 0-5. See Table S1 for ACE scoring details.

### Connectome Variate Associated with Cognitive Function and Psychopathology

In our previous study, we identified a brain functional connectivity-based dimension underlying individual differences in a wide range of cognitive functions and deviated behavioral/emotional functioning assessed using broad psychopathology measurements.^28^ This neural dimension is hypothesized to act as a resilience factor that can buffer the negative impact of ACEs on youth’s mental health.

The process to derive the connectome variate is detailed in Xiao et al., 2023.^28^ In brief, for each participant, a 20-min resting-state fMRI data was used to measure brain activities across 352 regions-of-interest (ROIs)^32^. The temporal synchrony between the ROIs was used to describe the individual’s functional brain connectome. A latent link between the functional connectome and the multidimensional behaviors was discovered using a machine learning approach called canonical correlation analysis (CCA), controlling for covariates in both brain and behavior datasets that may induce artificial brain-behavior associations. As this connectome variate (CV) exhibits positive correlations with cognitive function and negative correlations with psychopathology, we hypothesized that it acts as a potential neural index for resilience in the current study. See Figure S1 for more detailed description of the CV.

### Clinical Diagnoses for Mental Disorders

Youth psychiatric diagnoses were assessed using the computerized-version of the Kiddie Schedule for Affective Disorders and Schizophrenia for DSM-5 (KSADS-5), a psychometrically validated semi-structured psychiatric interview. ^33,34^ The KSADS-5 was administrated biannually from baseline. In the current study, 16 parent-reported KSADS-5 diagnoses at baseline and 2-year follow-up were included for analyses.

### Statistical Analysis

To confirm that ACEs are a transdiagnostic risk for mental disorders in the current cohort, we first grouped the participants based on their KSADS-5 diagnoses and compared the ACE score of each diagnostic group to the group without any diagnosis using Welch’s t-test. P values were corrected for multiple comparisons using the Benjamini-Hochberg false discovery rate (FDR) method ^35^. We then tested the relationship between the accumulative number of co-occurring KSADS disorders and the ACE score at baseline and year-2 using Pearson’s correlation.

We hypothesized that the association between ACEs and general psychopathology risk would be moderated by the CV score. We conducted three analyses to test this hypothesis.

First, we tested whether any associations between ACEs and KSADS Diagnoses were moderated by the baseline CV score, using the linear models: (# of KSADS Diagnoses)_baseline_ ∼ ACE_baseline_ * CV_baseline_ + ACE_baseline_ + CV_baseline_+ sex + race + age (Figure 3A), and (# of KSADS Diagnoses)_year2_ ∼ ACE_year2_ * CV_baseline_ + ACE_year2_ + CV_baseline_ + sex + race + age (Figure 4A). To investigate whether any such moderation effects were specific to the *Deprivation* and *Threat* subdimensions, the above analyses were repeated replacing the ACE total score with the individual sub-dimension scores.

**Figure 2.**
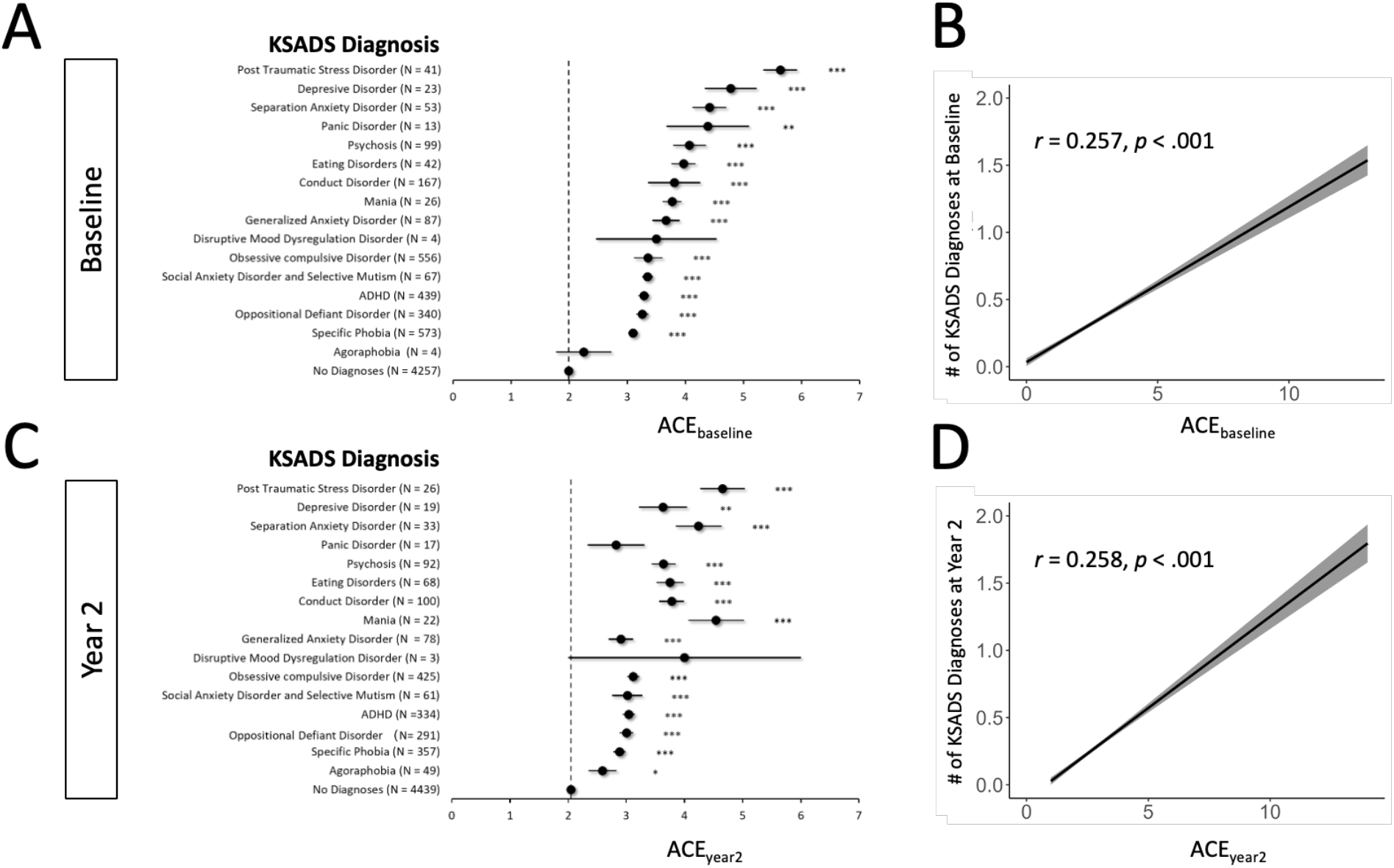
Relationship between ACEs and the mental health disorders, at baseline and 2-year follow up. A) Relationship between the occurrence of mental disorders and ACEs at baseline. B) Relationship between the accumulative occurrence across disorders and ACEs at baseline. C) Relationship between the occurrence of mental disorders and ACEs at 2-year follow up. D) Relationship between the accumulative occurrence across disorders and ACEs at 2-year follow up. In (A) and (C) error bars indicate the standard error. FDR-adjusted p-values: * p < 0.05, ** p < 0.01, *** p < 0.001. In (B) and (D) The black lines show the regression result of ‘# of KSADS Diagnoses ∼ ACE’ at baseline and year 2, respectively. The gray shadows show the 95% confidence interval of the regression result.

**Figure 3.**
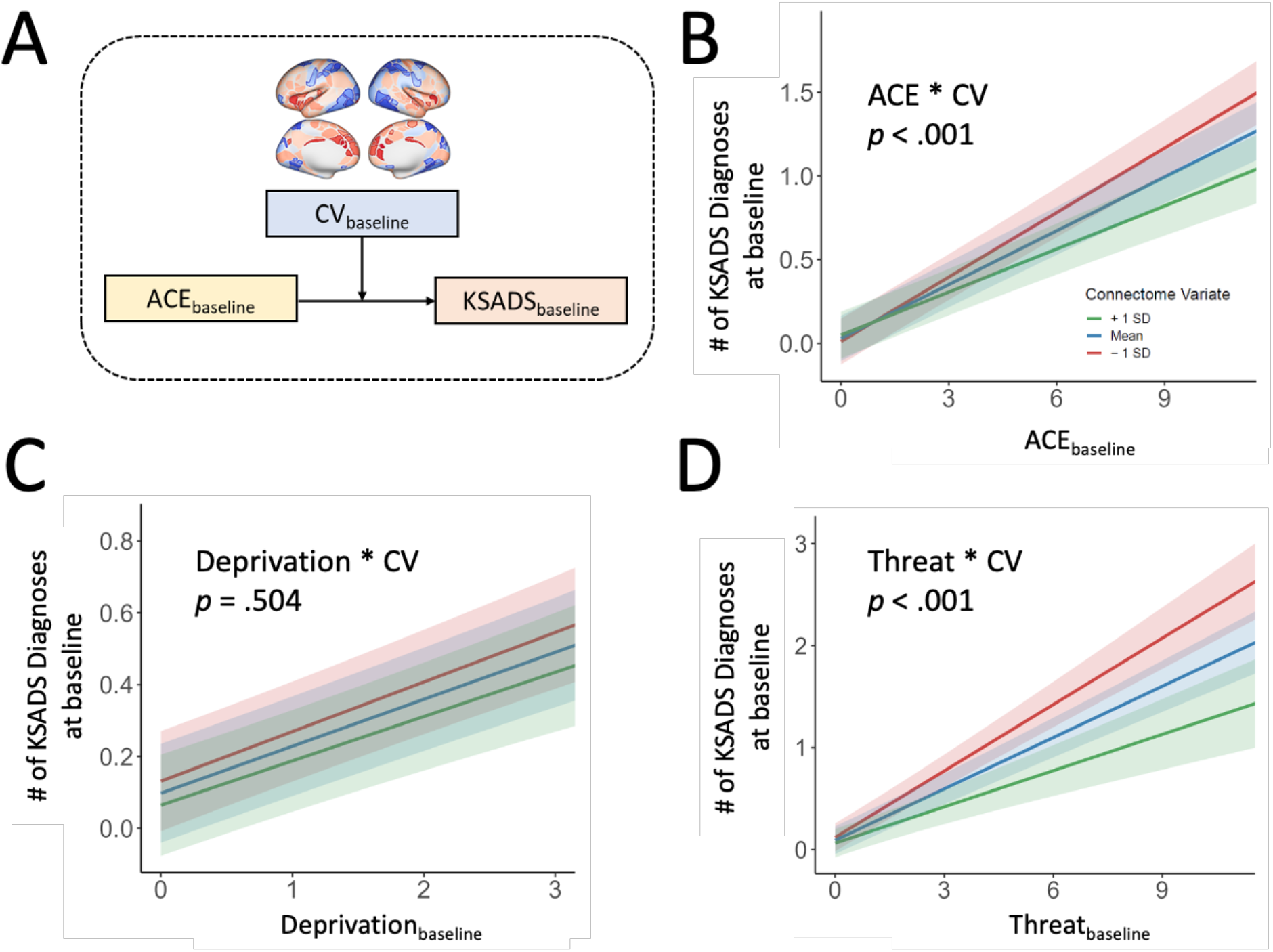
Moderation effect of the connectome variate on the association between baseline ACEs and the number of baseline KSADS diagnoses. A) Scheme of the moderation model. B-D) Moderation effect of CV on the association between the number of baseline KSADS diagnoses and ACEs, *Deprivation,* and *Threat*, respectively, at baseline. Moderation graphs show the model fit and 95% CI for the mean CV and the mean±1 SD.

**Figure 4.**
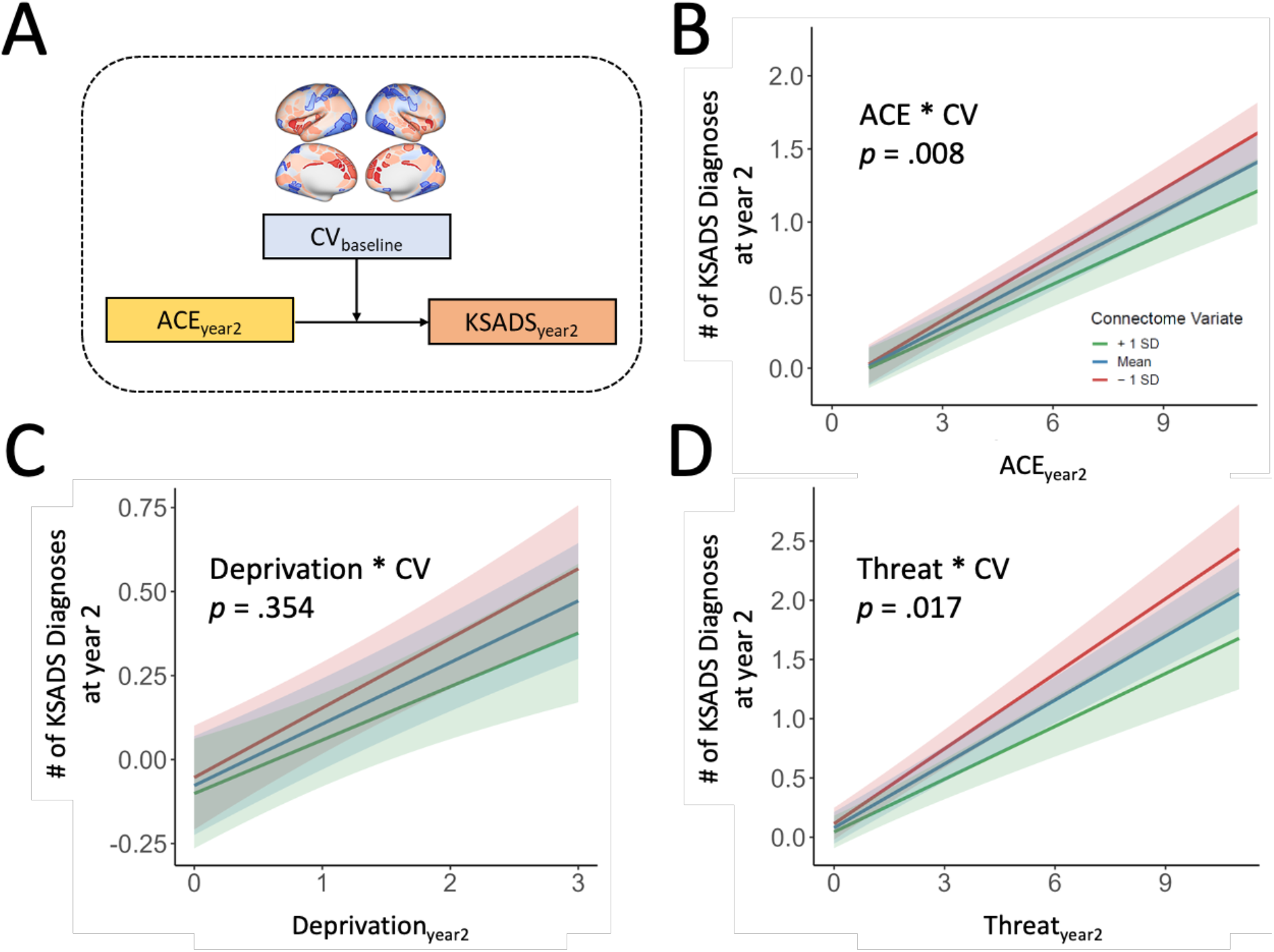
Moderation effect of the connectome variate on the association between year-2 ACEs and the number of year-2 KSADS diagnoses. A) Scheme of the moderation model. B-D) Moderation effect of CV on the association between the number of year-2 KSADS diagnoses and ACEs, *Deprivation,* and *Threat*, respectively, at the year-2 follow-up. Moderation graphs show the model fit and 95% CI for the mean CV and the mean±1 SD.

Second, to test whether the baseline CV modulated the changes in ACEs and the in the number of diagnoses from baseline to 2-year follow-up, we modified the year-2 moderation model by including the ACEs and number of KSADS diagnoses at baseline as covariates: (# of KSADS Diagnoses)_year2_ ∼ ACE_year2_ * CV_baseline_ + ACE_year2_ + ACE_baseline_ + (# of KSADS Diagnoses)_baseline_ + sex + race + age (Figure 5A).

**Figure 5.**
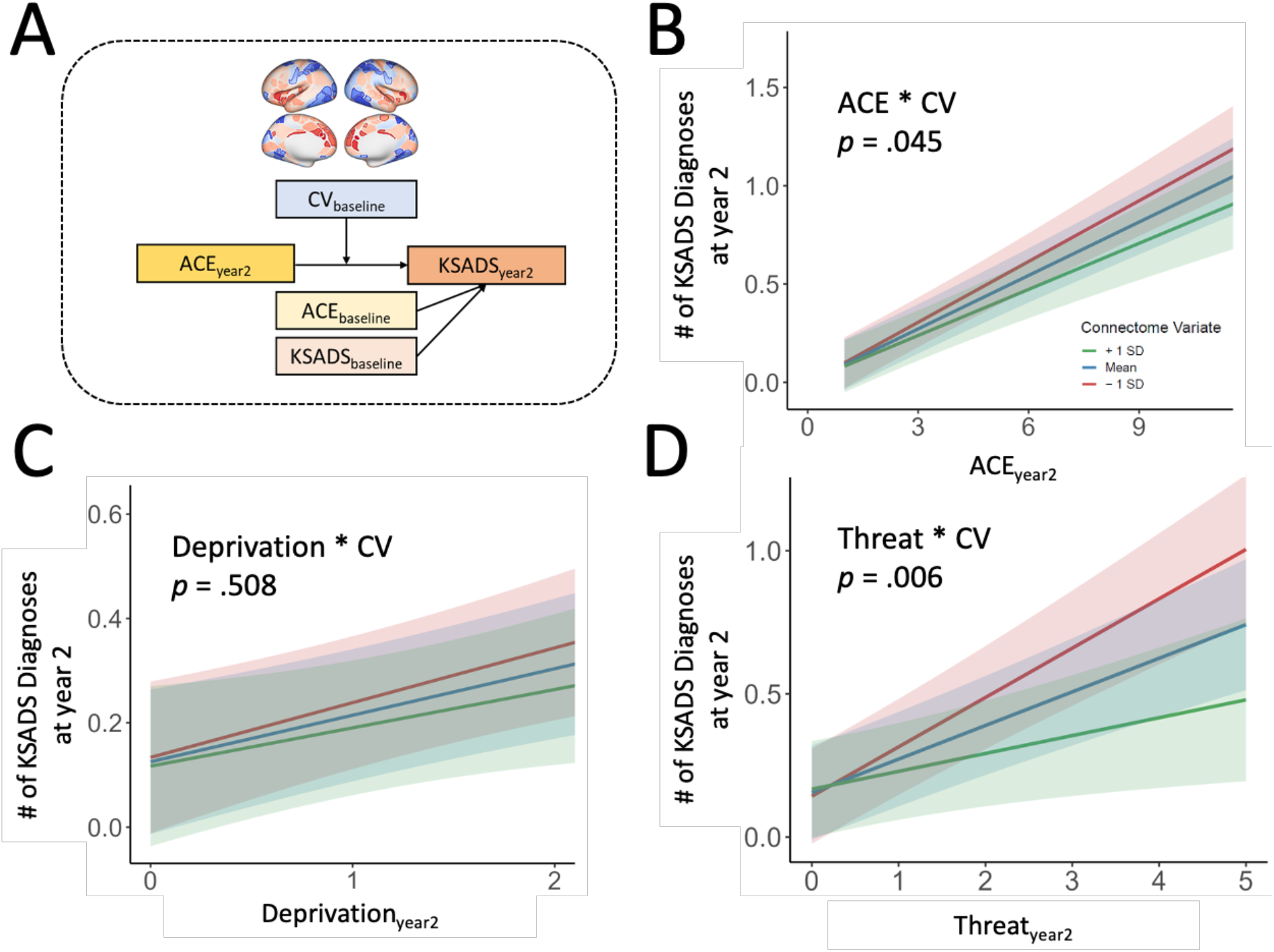
Moderation effect of the connectome variate on the association between changes in ACEs and in number of KSADS diagnoses from baseline to year-2 follow-up. A) Scheme of the moderation model. B) Moderation effect of CV on the association between the number of KSADS diagnoses and ACEs at Year 2, controlling for these two assessments at baseline. (C) and (D) Modulation effect for *Deprivation* and *Threat*, respectively, at year 2. Moderation graphs show the model fit and 95% CI for the mean CV and the mean±1 SD.

Finally, to test whether the above moderation effect depend on sex groups, we modified this model by adding sex as the secondary moderator: (# of KSADS Diagnoses)_year2_ ∼ sex * ACE_year2_ * CV_baseline_ + ACE_year2_ + CV_baseline_ ACE_baseline_ + (# of KSADS Diagnoses)_baseline_ + race + age (Figure 6A). The above ACE_year2_ * CV_baseline_ was further tested separately for the female and male groups.

The linear model analyses were conducted using the ‘lm4’ and R version 4.3.3.

## Results

### ACE as Transdiagnostic Risks for Mental Disorders

At both baseline and year-2 follow-up, most of the groups with KSADS diagnoses showed significantly higher ACE scores than the group with no current diagnosis (FDR-adjusted *p* < .05, Figures 2A and 2C). It further showed significant correlations between the ACE score and number of co-occurring disorders at baseline (Pearson’s *r* = 0.257, *p* < .001, Figure 2B) and year-2 follow-up (Pearson’s *r* = .258, *p* < .001, Figure 2D).

### Connectome Variate Moderates the Relationship Between Life-long ACEs and Psychiatric Diagnoses at Baseline

As shown in Figure 3B, CV significantly moderated the relationship between baseline ACE and KSADS diagnoses. (Interaction: ACE_baseline_ * CV, B = -0.021, SE = 0.005, *t* = -0.131, *p* < .001). Post-hoc analyses revealed that such moderation effect was significant on the relationship between KSADS and *Threat* (Interaction: *Threat*_baseline_ * CV, B = -0.049, SE = 0.012, *t* = -4.04, *p* < .001), but not on the relationship between KSADS and *Deprivation* (Interaction: *Deprivation*_baseline_ * CV, B = -0.007, SE = 0.011, *t* = -0.668, *p* = .504).

### Connectome Variate Moderates the Relationship Between Life-long ACEs and Psychiatric Diagnoses at Two-year Follow-up

Baseline CV significantly moderated the relationship between year-2 ACEs and KSADS diagnoses (Interaction: ACE_year2_ * CV, B = -0.018, SE = 0.006, *t* = -2.660, *p* = .008, N = 6327, Figure 4B). The moderation effect of CV was significant on the relationship between KSADS and *Threat* (Interaction: *Threat*_year2_ * CV, B = -0.032, SE = 0.013, *t* = -2.382, *p* < .001, Figure 4C), but not on the relationship between KSADS and *Deprivation* (Interaction: *Deprivation*_year2_ * CV, B = -0.024, SE = 0.026, *t* = -0.927, *p* = .354, Figure 4D).

### Connectome Variate Moderates the Relationship Between Recent ACEs and Development of Mental Disorders during the Two-Year Follow-Up Period

The baseline CV significantly moderated the relationship between changes in ACEs and in number of diagnoses from baseline to 2-year follow-up (Interaction: ACE_year2_ * CV, B = -0.012, SE = 0.006, *t* = -1.999, *p* = .045, shown in Figure 5B). As shown in Figures 4C and 4D, the moderation effect of CV was significant on the relationship between KSADS and *Threat* (Interaction: *Threat*_year2_ * CV, B = -0.033, SE = 0.011, *t* = -2.761, *p* = .006), but not on the relationship between KSADS and *Deprivation* (Interaction: *Threat*_year2_ * CV, B = -0.016, SE = 0.024, *t* = -0.661, *p* = .508).

### Sex Difference in the Prospective Moderation Effects of Connectome Variate

In a post-hoc analysis we examined whether the previous moderation effect depends on sex. We created a linear regression model to test for such a moderation effect (Figure S2A). The model revealed that the moderation effect of CV significantly depended on sex (Interaction: Threat_year2_ * CV * Sex, B = 0.05, SE = 0.237, *t* = 2.168, *p* = .03), with significant CV moderation effect in females (Interaction: Threat_year2_ * CV, B = -0.557, SE = 0.017, *t* = -3.368, *p* < .001, Figure S2B), but not males (Interaction: Threat_year2_ * CV, B = -0.001, SE = 0.017, *t* = -0.076, *p* = .940, Figure S2C). And such a sex-dependency was not observed in baseline (Interaction: Threat_baseline_ * CV * Sex, B = 0.058, SE = 0.010, *t* = 0.556, *p* = .58, Figure S3).

## Discussion

Resilience is a multi-disciplinary construct including factors contributing to one’s ability to maintain or regain mental health despite adversity experiences. Understanding stress-related disorders through the lens of what factors promote health was recently highlighted as a strategy to fill knowledge gaps that have remained unanswered by the traditional strategy of focusing on pathology ^16^. From a cohort of 9-10-year-old U.S. preadolescents, we have recently identified a functional connectome variate (CV) with significant loading on a broad system including frontoparietal cortices and subcortical nuclei. As the brain CV shows a robust positive association with better cognitive function and a negative association with transdiagnostic psychopathology, it was hypothesized in the current study as a potential neural marker for youth resilience against the risk of developing ACE-related psychiatric disorders. Leveraging the longitudinal design of the ABCD study, we confirmed that lifetime ACEs increased the likelihood of transdiagnostic psychopathology during the early stage of adolescence. We then discovered that the brain CV moderated the relationship between ACEs and transdiagnostic psychopathology such that individuals with higher CV scores had fewer current and 2-year follow-up KSADS diagnoses. In post-hoc analyses, we observed that the moderation effect of CV was specific to ACE subdimension of *threat* and specific to females.

Identifying neural markers subserving adolescents’ resilience prior to the ACE-related psychopathology is essential for protective interventions. However, in traditional resilience research, biological markers of individual’s resilience are rarely assessed before traumatic experiences like ACEs, making it difficult to clarify how the candidate biomarkers contribute to psychopathological consequences of ACEs ^16^. For example, a neural index associated with psychopathological response after ACEs can either predispose one’s resiliency adaptation with ACEs or express the acquired neural changes induced by the ACEs.^36^ Only recently have studies validated neuroimaging markers, via longitudinal designs, that predict ones’ resilience before exposure to life stressors and developing into relevant psychiatric symptoms ^19,36–38^. These identified resilience markers include attenuated activity in limbic regions such as the amygdala ^38^ and locus coeruleus ^37^, in response to stressful stimuli, and increased recruitment and activation of prefrontal brain regions involved in emotion regulation during emotional control tasks ^19,36^. Our finding that the brain CV moderated the relationship between ACEs and prospective KSADS diagnoses (two-year follow up) add to this evidence and further demonstrated that the adolescent brain connectome buffers the risk of future psychiatric disorders in youth who experienced ACEs. Given that the brain CV shows a high association with broad cognitive functions and carries significant loadings in a broad system consisting of frontoparietal cortices and subcortical nuclei, the CV may reflect the interplay of the two systems allowing for efficient emotion regulation, which warrants further investigation. Finally, as resting-state functional connectivity is reliable and suitable for repeated measures and has proved feasible for early stages of development such as school-age children ^39^ and even infants ^40^, the functional connectivity derived marker holds potential for tracking how resilience is cultivated before the onset of psychiatric disorders in adolescence.

ACEs have been proposed as a transdiagnostic risk factor ^5^. Our observation in the current study confirmed this notion in this cohort of 9–12-year-old U.S. youth, showing that accumulative adverse experiences were associated with heightened morbidity across a broad range of psychiatric disorders. According to the recent dimensional model of adversity, *threat* and *deprivation* have been hypothesized as distinct subdimensions that increase risk for psychiatric disorders via varied neurobiological pathways ^41^. This theory is based on evidence from developmental neuroscience research on both preclinical and clinical models, which shows the two subdimensions of ACEs are associated with structural and functional alternations in distinct brain systems through development ^42^. Our finding added to the evidence of the dimensional model in the brain CV showed dissociative moderation effects on the two subdimensions of ACE, significant for *threat* but not *deprivation*. Brain regions involved in stress response and regulation also exhibit pronounced loadings of the brain CV, including subcortical regions generating emotional response to stress, where glucocorticoid receptors are highly expressed ^43^, as well as ACC and insula which participate in fear inhibition ^44^. Such spatial overlap of stress and emotion regulation regions with high loadings in the brain CV may account for the CV’s protective effect to *threat.* Finally, the capability of the brain CV to prospectively predict youths’ resilience to *threat* versus *deprivation* may potentially provide a neural marker for identifying at-risk population and for informing precise intervention at early stage of ACE exposure.

Sex differences in resilience are of interest, as females generally show higher prevalence of stress related disorders ^45,46^. Though CV was slightly lower in females relative to males as shown in our previous study ^28^, the current study showed a CV moderation on the ACE-psychopathology association specifically in females. Additionally, such sex difference emerges between 9-10 to 11-12. Other studies have observed similar sex differences in the stress coping style. In adults, females tend to use more emotion-focused coping strategies, including emotion regulation and searching for social support ^47^. The growth of sex-specific tendency of using active coping strategies emerges during adolescence ^48,49^, and the emotion regulation capacity has also been found to predict life satisfaction specifically for female adolescents ^50^. If the brain CV serves as an intrinsic resource supporting the emotion regulation-based coping, as we speculate, its moderation effects on the ACE-disorder association could be pronounced in the population that relying more on such strategies. Preclinical research has also indicated that ACEs may affect psychopathology through sex-dependent mechanisms.^51^ Further investigations are warranted to specify the sociopsychological and/or biological factors that determines who may benefit more from higher CV, and how the benefits manifest for youth at different developmental stages, which may inform precise interventions for promoting youth resilience.

In conclusion, these findings provide preliminary evidence for a connectome-based resilience marker and suggest that functional connectivity strength in a broad system including frontal-parietal cortices and subcortical nuclei relevant to cognitive control may protect preadolescents who have experienced lifetime ACEs—especially females and those experiencing threat-related ACEs—from developing transdiagnostic psychopathology.

## Data Availability

All data produced in the present study are available upon reasonable request to the authors.

## Acknowledgments

This research was supported by the Intramural Research Program of the National Institute on Drug Abuse, National Institutes of Health.

Data used in the preparation of this article were obtained from the Adolescent Brain Cognitive Development (ABCD) Study (https://abcdstudy.org), held in the NIMH Data Archive (NDA). This is a multisite, longitudinal study designed to recruit more than 10,000 children aged 9-10 years and follow them over 10 years into early adulthood. The ABCD Study is supported by the National Institutes of Health and additional federal partners under award numbers U01DA041048, U01DA050989, U01DA051016, U01DA041022, U01DA051018, U01DA051037, U01DA050987, U01DA041174, U01DA041106, U01DA041117, U01DA041028, U01DA041134, U01DA050988, U01DA051039, U01DA041156, U01DA041025, U01DA041120, U01DA051038, U01DA041148, U01DA041093, U01DA041089, U24DA041123, U24DA041147. A full list of supporters is available at https://abcdstudy.org/federal-partners.html. A listing of participating sites and a complete listing of the study investigators can be found at https://abcdstudy.org/consortium_members/. ABCD consortium investigators designed and implemented the study and/or provided data but did not necessarily participate in analysis or writing of this report. This manuscript reflects the views of the authors and may not reflect the opinions or views of the NIH or ABCD consortium investigators.

The ABCD data repository grows and changes over time. The ABCD data used in this report came from NDA Study 901. DOIs can be found at DOI 10.15154/1519007.

This work utilized the computational resources of the NIH HPC Biowulf cluster (http://hpc.nih.gov).

