## Supplementary Materials for "Connectome-based Brain Marker Moderates the Relationship between Childhood Adversity and Transdiagnostic Psychopathology during Early Adolescence"

### *Supplementary Methods*

#### Scoring of ACE

Table S1 lists the scoring ACEs in the ABCD dataset, following the scheme of Stinson et al 2021[2]. Life events included in the ACE category were: Emotion Abuse, Physical Abuse, Sexual Abuse, Domestic Violence, Traumatic Grief, Community Violence, Nature Disaster, Fire, Experience of War Zone, Experience of Terrorism, Car Accident, Other Significant Accident, Bullying, Physical Neglect, Emotional Neglect, Household Substance Use, Mental Illness in Household, Family Member Involved in Criminal Justice System, Parental Separation or Divorce, Racial or Ethnic Discrimination, and Financial Adversity. According to Sheridan & McLaughlin [3], two subdimensions, *Threat* and *Deprivation*, were derived from the ACE events. *Threat* involved the experiences that were unexpected and could do harm to or threaten one's physical integrity. The *Threat* dimension score included Emotion Abuse, Physical Abuse, Sexual Abuse, Domestic Violence, Traumatic Grief, Community Violence, Nature Disaster, Fire, Experience of War Zone, Experience of Terrorism, Car Accident, Other Significant Accident, and Bullying. The *Threat* score ranged between 0-13. *Deprivation* involved the absence of expected social and cognitive environment for the children. In the current study, the *Deprivation* dimension score included Physical Neglect, Emotional Neglect, Parental Separation or Divorce, Racial or Ethnic Discrimination, and Financial Adversity. The *Deprivation* score ranged between 0-5.

Some questionnaires used for calculating ACE were missing at baseline or year-2. In those cases, year-1 data were used. The Perceived Discrimination Survey and Adverse Life Events Survey were collected yearly since year 1; therefore baseline data for the two surveys are unavailable. We used the year-1 data for the baseline assessment. The Children's Report of Parental Behavioral

Inventory was collected at baseline and changed to bi-annually assessment since year 1; therefore year-2 data are not available. We used the year-1 data for the year-2 assessment.

#### *Supplementary Figures and Tables*

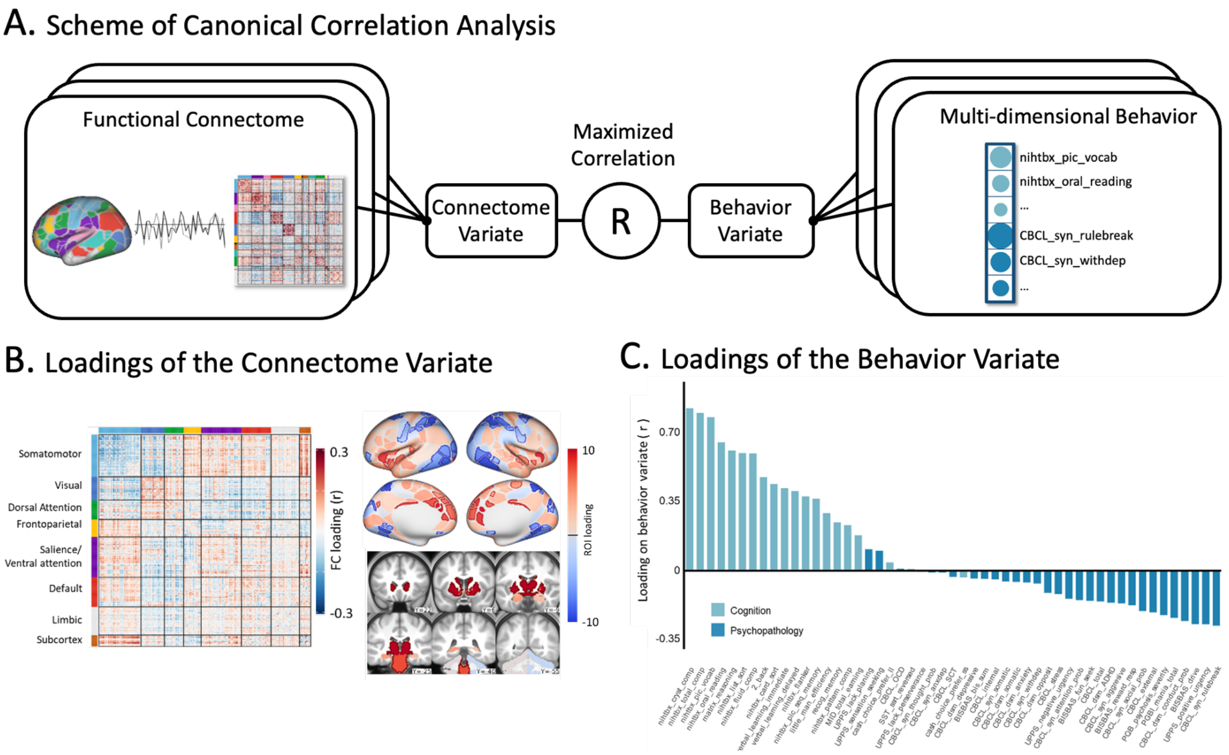


**Figure S1 The connectome variate associated with cognition and mental health. A) Scheme of canonical correlation analysis (CCA).** CCA was used to detect latent association between the two data sets of functional connectome and multi-dimensional behavior assessments including cognitive test and psychopathological assessments. **B) Loading of the connectome variate.** The left panel shows the functional connectivity-wise loading. The right panel shows the ROI-sum of loading, borderlines highlight ROIs showing significant positive and negative loading. **C) Loading of the behavior variate.** Figure is produced from data reported in our previous study of Xiao et al, 2023 [1].

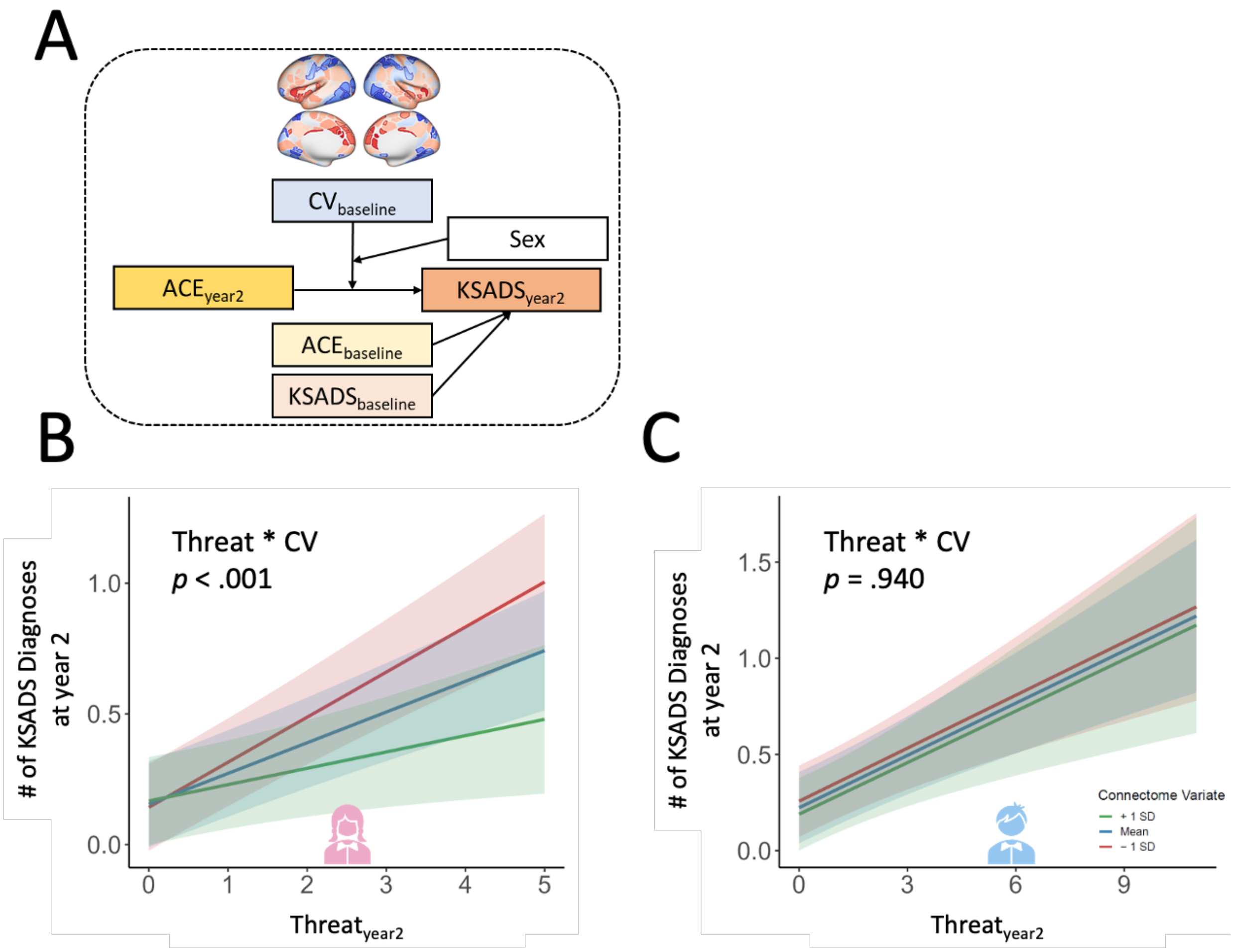


**Figure S2. Sex difference in the modulation of baseline CV on the association between the change in Threat and in the number of KSADS diagnoses from baseline to year-2 follow-up. B-C) Moderation effect of CV on the association between *Threat* and number of KSADS diagnoses at year-2 follow-up in females and males, respectively, with controlling for total ACE and the number of KSADS diagnoses at baseline as covariates.** Moderation graphs show the model fit and 95% CI for the mean CV and the mean±1 SD.

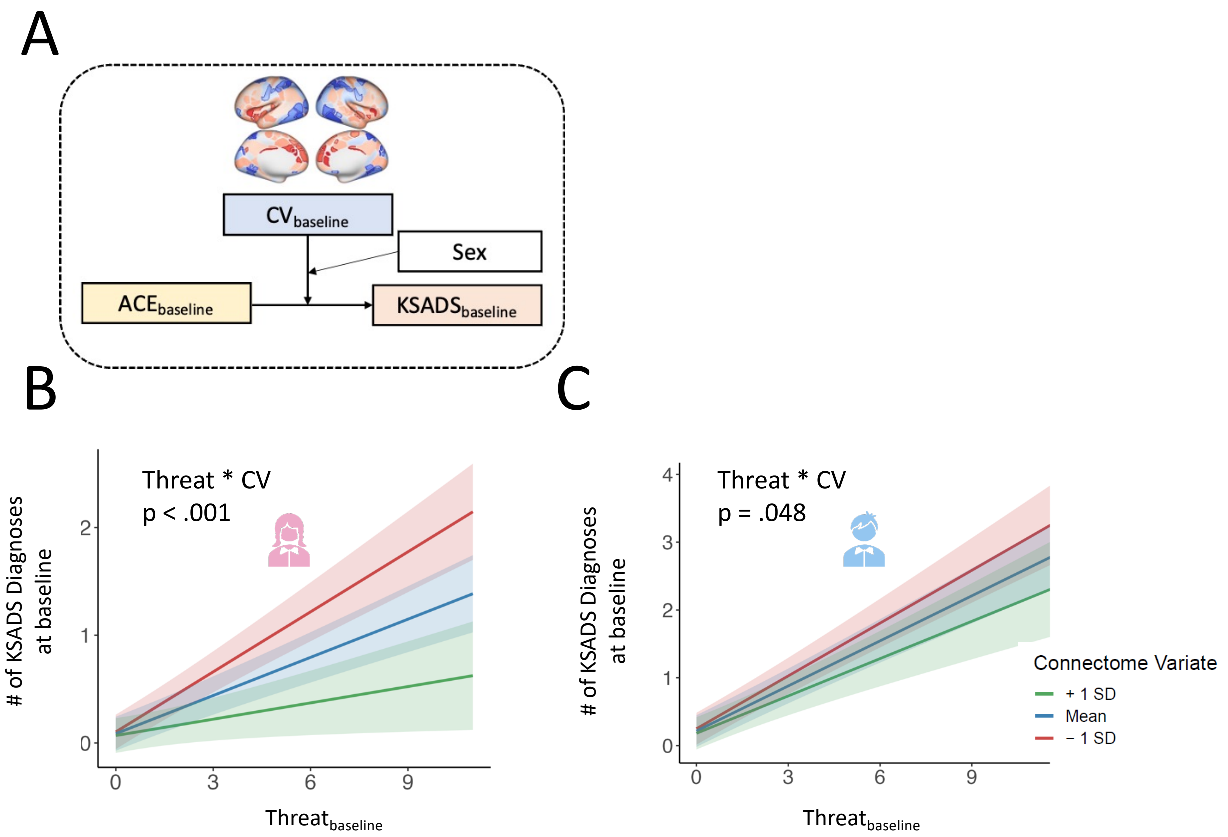


**Figure S3. Sex difference in the modulation of baseline CV on the association between Threat and mental disorder diagnoses at baseline. A) Scheme of the moderation model. B-C) Moderation effect of CV on the association between *Threat* and number of KSADS diagnoses at baseline in females and males, respectively.** Moderation graphs show the model fit and 95% CI for the mean CV and the mean±1 SD.

**Table S1 Scoring Scheme for Adverse Childhood Experience**

| Subdimension | Measurement | Criteria | Item-wise criteria | Item |
|---|---|---|---|---|
| Threat | Emotional Abuse | score 1 if criteria are met for any of the questions | yes = 1; no = 0 | ksads_ptsd_raw_764_p |
| | | | yes = 1; no = 0 | ksads_ptsd_raw_765_p |
| | Physical Abuse | score 1 if criteria are met for any of the questions | yes = 1; no = 0 | ksads_ptsd_raw_761_p |
| | | | yes = 1; no = 0 | ksads_ptsd_raw_762_p |
| | Sexual Abuse | score 1 if criteria are met for any of the questions | yes = 1; no = 0 | ksads_ptsd_raw_767_p |
| | | | yes = 1; no = 0 | ksads_ptsd_raw_768_p |
| | | | yes = 1; no = 0 | ksads_ptsd_raw_769_p |
| | Domestic Violence | score 1 if criteria are meet | yes = 1; no = 0 | ksads_ptsd_raw_766_p |
| | Traumatic Grief | score 1 if criteria are meet | yes = 1; no = 0 | ksads_ptsd_raw_770_p |
| | Community Violence | score 1 if criteria are meet | yes = 1; no = 0 | ksads_ptsd_raw_760_p |
| | Natural Disaster | score 1 if criteria are meet | yes = 1; no = 0 | ksads_ptsd_raw_757_p |
| | Fire | score 1 if criteria are meet | yes = 1; no = 0 | ksads_ptsd_raw_756_p |
| | Experience of War Zone | score 1 if criteria are meet | yes = 1; no = 0 | ksads_ptsd_raw_759_p |
| | Experience of Terrorism | score 1 if criteria are meet | yes = 1; no = 0 | ksads_ptsd_raw_758_p |
| | Car Accident | score 1 if criteria are meet | yes = 1; no = 0 | ksads_ptsd_raw_754_p |
| | Other Significant Accident | score 1 if criteria are meet | yes = 1; no = 0 | ksads_ptsd_raw_755_p |
| | Bullying | score 1 if criteria are meet | yes = 1; no = 0 | ksads_bully_raw_26 |
| Deprivation | Physical Negnect | score 1 if criteria are met for any of the questions | 1-2=1 ; 3-5 = 0 | parent_monitor_q1_y |
| | | | 1-2=1 ; 3-5 = 0 | parent_monitor_q2_y |
| | | | 1-2=1 ; 3-5 = 0 | parent_monitor_q4_y |
| | Emotional Neglect | Score 1 if >2 out the 5 questions meet criteria. | 1 = 0; 2-3 = 1 | crpbi_parent1_y |
| | | | 1 = 0; 2-3 = 1 | crpbi_parent2_y |
| | | | 1 = 0; 2-3 = 1 | crpbi_parent3_y |
| | | | 1 = 0; 2-3 = 1 | crpbi_parent4_y |
| | | | 1 = 0; 2-3 = 1 | crpbi_parent5_y |
| | Racial or Ethnic Discrimination | score 1 if both criteria are met | yes = 1; No = 0 | dim_yesno_q1 |
| | | | 1-2 = 0 (almost never or rarely); 3-5 = 1 (sometimes to very often) | dim_matrix_q1 |
| | | | | dim_matrix_q2 |
| | | | | dim_matrix_q3 |
| | Parental Separation or Divorce | score 1 if any of the questions meet criteria | 1 = 0; 2-6 = 1 | demo_prnt_marital_v2_l |
| | | | yes = 1; no = 0 | ple_separ_p |
| | | | yes = 1; no = 0 | ple_separ_y |
| | Financial Adversity | score 1 if any of the questions meet criteria | yes = 1; no = 0 | demo_fam_exp1_v2_l |
| | | | yes = 1; no = 0 | demo_fam_exp3_v2_l |
| | | | yes = 1; no = 0 | demo_fam_exp4_v2_l |
| | | | yes = 1; no = 0 | demo_fam_exp5_v2_l |
| | | | yes = 1; no = 0 | demo_fam_exp6_v2_l |
| Others | Household Substance Use | score 1 if any of the questions meet criteria | yes = 1; no = 0 | famhx_ss_fath_prob_dg_p |
| | | | yes = 1; no = 0 | famhx_ss_moth_prob_dg_p |
| | | | 0 = 0; 1-2 = 1 | asr_q06_p |
| | | | 0 = 0; 1-2 = 1 | asr_q90_p |
| | | | yes = 1; no = 0 | famhx_ss_fulsiby1_prob_dg_p |
| | | | yes = 1; no = 0 | famhx_ss_fulsiby2_prob_dg_p |
| | | | yes = 1; no = 0 | famhx_ss_fulsiby3_prob_dg_p |
| | | | yes = 1; no = 0 | famhx_ss_fulsiby4_prob_dg_p |
| | | | yes = 1; no = 0 | famhx_ss_fulsiby5_prob_dg_p |
| | | | yes = 1; no = 0 | famhx_ss_fulsibo1_prob_dg_p |

| | | | | |
|---|---|---|---|---|
| | | | -yes = 1; no = 0 | famhx_ss_fulsibo2_prob_dg_p |
| | | | yes = 1; no = 0 | famhx_ss_fulsibo3_prob_dg_p |
| | | | yes = 1; no = 0 | famhx_ss_fulsibo4_prob_dg_p |
| | | | yes = 1; no = 0 | famhx_ss_hlfsiby1_prob_dg_p |
| | | | yes = 1; no = 0 | famhx_ss_hlfsiby2_prob_dg_p |
| | | | yes = 1; no = 0 | famhx_ss_hlfsiby3_prob_dg_p |
| | | | yes = 1; no = 0 | famhx_ss_hlfsiby4_prob_dg_p |
| | | | yes = 1; no = 0 | famhx_ss_hlfsiby5_prob_dg_p |
| | | | yes = 1; no = 0 | famhx_ss_hlfsibo1_prob_dg_p |
| | | | yes = 1; no = 0 | famhx_ss_hlfsibo2_prob_dg_p |
| | | | yes = 1; no = 0 | famhx_ss_hlfsibo3_prob_dg_p |
| | | | yes = 1; no = 0 | famhx_ss_hlfsibo4_prob_dg_p |
| | | | yes = 1; no = 0 | famhx_ss_hlfsibo5_prob_dg_p |
| | | | yes = 1; no = 0 | famhx_ss_fulsibo5_prob_dg_p |
| | | | yes = 1; no = 0 | ple_sud_p |
| | | | yes = 1; no = 0 | ple_sud_y |
| | Mental Illness in Household | score 1 if any of the questions meet criteria | yes = 1; no = 0 | famhx_ss_fulsiby1_prob_dprs_p |
| | | | yes = 1; no = 0 | famhx_ss_fulsiby2_prob_dprs_p |
| | | | yes = 1; no = 0 | famhx_ss_fulsiby3_prob_dprs_p |
| | | | yes = 1; no = 0 | famhx_ss_fulsiby4_prob_dprs_p |
| | | | yes = 1; no = 0 | famhx_ss_fulsiby5_prob_dprs_p |
| | | | yes = 1; no = 0 | famhx_ss_fulsibo1_prob_dprs_p |
| | | | yes = 1; no = 0 | famhx_ss_fulsibo2_prob_dprs_p |
| | | | yes = 1; no = 0 | famhx_ss_fulsibo3_prob_dprs_p |
| | | | yes = 1; no = 0 | famhx_ss_fulsibo4_prob_dprs_p |
| | | | yes = 1; no = 0 | famhx_ss_fulsibo5_prob_dprs_p |
| | | | yes = 1; no = 0 | famhx_ss_hlfsiby1_prob_dprs_p |
| | | | yes = 1; no = 0 | famhx_ss_hlfsiby2_prob_dprs_p |
| | | | yes = 1; no = 0 | famhx_ss_hlfsiby3_prob_dprs_p |
| | | | yes = 1; no = 0 | famhx_ss_hlfsiby4_prob_dprs_p |
| | | | yes = 1; no = 0 | famhx_ss_hlfsiby5_prob_dprs_p |
| | | | yes = 1; no = 0 | famhx_ss_hlfsibo1_prob_dprs_p |
| | | | yes = 1; no = 0 | famhx_ss_hlfsibo2_prob_dprs_p |
| | | | yes = 1; no = 0 | famhx_ss_hlfsibo3_prob_dprs_p |
| | | | yes = 1; no = 0 | famhx_ss_hlfsibo4_prob_dprs_p |
| | | | yes = 1; no = 0 | famhx_ss_hlfsibo5_prob_dprs_p |
| | | | yes = 1; no = 0 | famhx_ss_momdad_dprs_p |
| | | | yes = 1; no = 0 | famhx_ss_fulsiby1_prob_ma_p |
| | | | yes = 1; no = 0 | famhx_ss_fulsiby2_prob_ma_p |
| | | | yes = 1; no = 0 | famhx_ss_fulsiby3_prob_ma_p |
| | | | yes = 1; no = 0 | famhx_ss_fulsiby4_prob_ma_p |
| | | | yes = 1; no = 0 | famhx_ss_fulsiby5_prob_ma_p |
| | | | yes = 1; no = 0 | famhx_ss_fulsibo1_prob_ma_p |
| | | | yes = 1; no = 0 | famhx_ss_fulsibo2_prob_ma_p |
| | | | yes = 1; no = 0 | famhx_ss_fulsibo3_prob_ma_p |
| | | | yes = 1; no = 0 | famhx_ss_fulsibo4_prob_ma_p |
| | | | yes = 1; no = 0 | famhx_ss_fulsibo5_prob_ma_p |
| | | | yes = 1; no = 0 | famhx_ss_hlfsiby1_prob_ma_p |
| | | | yes = 1; no = 0 | famhx_ss_hlfsiby2_prob_ma_p |
| | | | yes = 1; no = 0 | famhx_ss_hlfsiby3_prob_ma_p |
| | | | yes = 1; no = 0 | famhx_ss_hlfsiby4_prob_ma_p |
| | | | yes = 1; no = 0 | famhx_ss_hlfsiby5_prob_ma_p |
| | | | yes = 1; no = 0 | famhx_ss_hlfsibo1_prob_ma_p |
| | | | yes = 1; no = 0 | famhx_ss_hlfsibo2_prob_ma_p |
| | | | yes = 1; no = 0 | famhx_ss_hlfsibo3_prob_ma_p |
| | | | yes = 1; no = 0 | famhx_ss_hlfsibo4_prob_ma_p |
| | | | yes = 1; no = 0 | famhx_ss_hlfsibo5_prob_ma_p |
| | | | yes = 1; no = 0 | famhx_ss_momdad_ma_p |
| | | | yes = 1; no = 0 | famhx_ss_fulsiby1_prob_vs_p |
| | | | yes = 1; no = 0 | famhx_ss_fulsiby2_prob_vs_p |
| | | | yes = 1; no = 0 | famhx_ss_fulsiby3_prob_vs_p |
| | | | yes = 1; no = 0 | famhx_ss_fulsiby4_prob_vs_p |
| | | | yes = 1; no = 0 | famhx_ss_fulsiby5_prob_vs_p |

| | | | | |
|---|---|---|---|---|
| | | | yes = 1; no = 0 | famhx_ss_fulsibo1_prob_vs_p |
| | | | yes = 1; no = 0 | famhx_ss_fulsibo2_prob_vs_p |
| | | | yes = 1; no = 0 | famhx_ss_fulsibo3_prob_vs_p |
| | | | yes = 1; no = 0 | famhx_ss_fulsibo4_prob_vs_p |
| | | | yes = 1; no = 0 | famhx_ss_fulsibo5_prob_vs_p |
| | | | yes = 1; no = 0 | famhx_ss_hlfsiby1_prob_vs_p |
| | | | yes = 1; no = 0 | famhx_ss_hlfsiby2_prob_vs_p |
| | | | yes = 1; no = 0 | famhx_ss_hlfsiby3_prob_vs_p |
| | | | yes = 1; no = 0 | famhx_ss_hlfsiby4_prob_vs_p |
| | | | yes = 1; no = 0 | famhx_ss_hlfsiby5_prob_vs_p |
| | | | yes = 1; no = 0 | famhx_ss_hlfsibo1_prob_vs_p |
| | | | yes = 1; no = 0 | famhx_ss_hlfsibo2_prob_vs_p |
| | | | yes = 1; no = 0 | famhx_ss_hlfsibo3_prob_vs_p |
| | | | yes = 1; no = 0 | famhx_ss_hlfsibo4_prob_vs_p |
| | | | yes = 1; no = 0 | famhx_ss_hlfsibo5_prob_vs_p |
| | | | yes = 1; no = 0 | famhx_ss_momdad_vs_p |
| | | | yes = 1; no = 0 | famhx_ss_fulsiby1_prob_trb_p |
| | | | yes = 1; no = 0 | famhx_ss_fulsiby2_prob_trb_p |
| | | | yes = 1; no = 0 | famhx_ss_fulsiby3_prob_trb_p |
| | | | yes = 1; no = 0 | famhx_ss_fulsiby4_prob_trb_p |
| | | | yes = 1; no = 0 | famhx_ss_fulsiby5_prob_trb_p |
| | | | yes = 1; no = 0 | famhx_ss_fulsibo1_prob_trb_p |
| | | | yes = 1; no = 0 | famhx_ss_fulsibo2_prob_trb_p |
| | | | yes = 1; no = 0 | famhx_ss_fulsibo3_prob_trb_p |
| | | | yes = 1; no = 0 | famhx_ss_fulsibo4_prob_trb_p |
| | | | yes = 1; no = 0 | famhx_ss_fulsibo5_prob_trb_p |
| | | | yes = 1; no = 0 | famhx_ss_hlfsiby1_prob_trb_p |
| | | | yes = 1; no = 0 | famhx_ss_hlfsiby2_prob_trb_p |
| | | | yes = 1; no = 0 | famhx_ss_hlfsiby3_prob_trb_p |
| | | | yes = 1; no = 0 | famhx_ss_hlfsiby4_prob_trb_p |
| | | | yes = 1; no = 0 | famhx_ss_hlfsiby5_prob_trb_p |
| | | | yes = 1; no = 0 | famhx_ss_hlfsibo1_prob_trb_p |
| | | | yes = 1; no = 0 | famhx_ss_hlfsibo2_prob_trb_p |
| | | | yes = 1; no = 0 | famhx_ss_hlfsibo3_prob_trb_p |
| | | | yes = 1; no = 0 | famhx_ss_hlfsibo4_prob_trb_p |
| | | | yes = 1; no = 0 | famhx_ss_hlfsibo5_prob_trb_p |
| | | | yes = 1; no = 0 | famhx_ss_momdad_trb_p |
| | | | yes = 1; no = 0 | famhx_ss_fulsiby1_prob_nrv_p |
| | | | yes = 1; no = 0 | famhx_ss_fulsiby2_prob_nrv_p |
| | | | yes = 1; no = 0 | famhx_ss_fulsiby3_prob_nrv_p |
| | | | yes = 1; no = 0 | famhx_ss_fulsiby4_prob_nrv_p |
| | | | yes = 1; no = 0 | famhx_ss_fulsiby5_prob_nrv_p |
| | | | yes = 1; no = 0 | famhx_ss_fulsibo1_prob_nrv_p |
| | | | yes = 1; no = 0 | famhx_ss_fulsibo2_prob_nrv_p |
| | | | yes = 1; no = 0 | famhx_ss_fulsibo3_prob_nrv_p |
| | | | yes = 1; no = 0 | famhx_ss_fulsibo4_prob_nrv_p |
| | | | yes = 1; no = 0 | famhx_ss_fulsibo5_prob_nrv_p |
| | | | yes = 1; no = 0 | famhx_ss_hlfsiby1_prob_nrv_p |
| | | | yes = 1; no = 0 | famhx_ss_hlfsiby2_prob_nrv_p |
| | | | yes = 1; no = 0 | famhx_ss_hlfsiby3_prob_nrv_p |
| | | | yes = 1; no = 0 | famhx_ss_hlfsiby4_prob_nrv_p |
| | | | yes = 1; no = 0 | famhx_ss_hlfsiby5_prob_nrv_p |
| | | | yes = 1; no = 0 | famhx_ss_hlfsibo1_prob_nrv_p |
| | | | yes = 1; no = 0 | famhx_ss_hlfsibo2_prob_nrv_p |
| | | | yes = 1; no = 0 | famhx_ss_hlfsibo3_prob_nrv_p |
| | | | yes = 1; no = 0 | famhx_ss_hlfsibo4_prob_nrv_p |
| | | | yes = 1; no = 0 | famhx_ss_hlfsibo5_prob_nrv_p |
| | | | yes = 1; no = 0 | famhx_ss_momdad_nrv_p |
| | | | yes = 1; no = 0 | famhx_ss_fulsiby1_prob_prf_p |
| | | | yes = 1; no = 0 | famhx_ss_fulsiby2_prob_prf_p |
| | | | yes = 1; no = 0 | famhx_ss_fulsiby3_prob_prf_p |
| | | | yes = 1; no = 0 | famhx_ss_fulsiby4_prob_prf_p |
| | | | yes = 1; no = 0 | famhx_ss_fulsiby5_prob_prf_p |

| | | | | |
|---|---|---|---|---|
| | | | yes = 1; no = 0 | famhx_ss_fulsibo1_prob_prf_p |
| | | | yes = 1; no = 0 | famhx_ss_fulsibo2_prob_prf_p |
| | | | yes = 1; no = 0 | famhx_ss_fulsibo3_prob_prf_p |
| | | | yes = 1; no = 0 | famhx_ss_fulsibo4_prob_prf_p |
| | | | yes = 1; no = 0 | famhx_ss_fulsibo5_prob_prf_p |
| | | | yes = 1; no = 0 | famhx_ss_hlfsiby1_prob_prf_p |
| | | | yes = 1; no = 0 | famhx_ss_hlfsiby2_prob_prf_p |
| | | | yes = 1; no = 0 | famhx_ss_hlfsiby3_prob_prf_p |
| | | | yes = 1; no = 0 | famhx_ss_hlfsiby4_prob_prf_p |
| | | | yes = 1; no = 0 | famhx_ss_hlfsiby5_prob_prf_p |
| | | | yes = 1; no = 0 | famhx_ss_hlfsibo1_prob_prf_p |
| | | | yes = 1; no = 0 | famhx_ss_hlfsibo2_prob_prf_p |
| | | | yes = 1; no = 0 | famhx_ss_hlfsibo3_prob_prf_p |
| | | | yes = 1; no = 0 | famhx_ss_hlfsibo4_prob_prf_p |
| | | | yes = 1; no = 0 | famhx_ss_hlfsibo5_prob_prf_p |
| | | | yes = 1; no = 0 | famhx_ss_momdad_prf_p |
| | | | yes = 1; no = 0 | famhx_ss_fulsiby1_prob_hspd_p |
| | | | yes = 1; no = 0 | famhx_ss_fulsiby2_prob_hspd_p |
| | | | yes = 1; no = 0 | famhx_ss_fulsiby3_prob_hspd_p |
| | | | yes = 1; no = 0 | famhx_ss_fulsiby4_prob_hspd_p |
| | | | yes = 1; no = 0 | famhx_ss_fulsiby5_prob_hspd_p |
| | | | yes = 1; no = 0 | famhx_ss_fulsibo1_prob_hspd_p |
| | | | yes = 1; no = 0 | famhx_ss_fulsibo2_prob_hspd_p |
| | | | yes = 1; no = 0 | famhx_ss_fulsibo3_prob_hspd_p |
| | | | yes = 1; no = 0 | famhx_ss_fulsibo4_prob_hspd_p |
| | | | yes = 1; no = 0 | famhx_ss_fulsibo5_prob_hspd_p |
| | | | yes = 1; no = 0 | famhx_ss_hlfsiby1_prob_hspd_p |
| | | | yes = 1; no = 0 | famhx_ss_hlfsiby2_prob_hspd_p |
| | | | yes = 1; no = 0 | famhx_ss_hlfsiby3_prob_hspd_p |
| | | | yes = 1; no = 0 | famhx_ss_hlfsiby4_prob_hspd_p |
| | | | yes = 1; no = 0 | famhx_ss_hlfsiby5_prob_hspd_p |
| | | | yes = 1; no = 0 | famhx_ss_hlfsibo1_prob_hspd_p |
| | | | yes = 1; no = 0 | famhx_ss_hlfsibo2_prob_hspd_p |
| | | | yes = 1; no = 0 | famhx_ss_hlfsibo3_prob_hspd_p |
| | | | yes = 1; no = 0 | famhx_ss_hlfsibo4_prob_hspd_p |
| | | | yes = 1; no = 0 | famhx_ss_hlfsibo5_prob_hspd_p |
| | | | yes = 1; no = 0 | famhx_ss_momdad_hspd_p |
| | | | yes = 1; no = 0 | famhx_ss_fulsiby1_prob_scd_p |
| | | | yes = 1; no = 0 | famhx_ss_fulsiby2_prob_scd_p |
| | | | yes = 1; no = 0 | famhx_ss_fulsiby3_prob_scd_p |
| | | | yes = 1; no = 0 | famhx_ss_fulsiby4_prob_scd_p |
| | | | yes = 1; no = 0 | famhx_ss_fulsiby5_prob_scd_p |
| | | | yes = 1; no = 0 | famhx_ss_fulsibo1_prob_scd_p |
| | | | yes = 1; no = 0 | famhx_ss_fulsibo2_prob_scd_p |
| | | | yes = 1; no = 0 | famhx_ss_fulsibo3_prob_scd_p |
| | | | yes = 1; no = 0 | famhx_ss_fulsibo4_prob_scd_p |
| | | | yes = 1; no = 0 | famhx_ss_fulsibo5_prob_scd_p |
| | | | yes = 1; no = 0 | famhx_ss_hlfsiby1_prob_scd_p |
| | | | yes = 1; no = 0 | famhx_ss_hlfsiby2_prob_scd_p |
| | | | yes = 1; no = 0 | famhx_ss_hlfsiby3_prob_scd_p |
| | | | yes = 1; no = 0 | famhx_ss_hlfsiby4_prob_scd_p |
| | | | yes = 1; no = 0 | famhx_ss_hlfsiby5_prob_scd_p |
| | | | yes = 1; no = 0 | famhx_ss_hlfsibo1_prob_scd_p |
| | | | yes = 1; no = 0 | famhx_ss_hlfsibo2_prob_scd_p |
| | | | yes = 1; no = 0 | famhx_ss_hlfsibo3_prob_scd_p |
| | | | yes = 1; no = 0 | famhx_ss_hlfsibo4_prob_scd_p |
| | | | yes = 1; no = 0 | famhx_ss_hlfsibo5_prob_scd_p |
| | | | yes = 1; no = 0 | famhx_ss_momdad_scd_p |
| | | | t>63 = 1; t<=63 = 0; | asr_scr_depress_t |
| | | | t>63 = 1; t<=63 = 0; | asr_scr_anxdisord_t |
| | | | t>63 = 1; t<=63 = 0; | asr_scr_avoidant_t |
| | | | t>63 = 1; t<=63 = 0; | asr_scr_adhd_t |
| | | | t>63 = 1; t<=63 = 0; | asr_scr_antisocial_t |

| | | | | |
|---|---|---|---|---|
| | Family member involved in criminal justice system | score 1 if any of the questions meet criteria | yes = 1; no = 0 | ple_arrest_p |
| | | | yes = 1; no = 0 | ple_law_p |
| | | | yes = 1; no = 0 | ple_jail_p |
